# Discovery and Validation of Gut Microbiome Features Associated with Dietary Patterns in U.S. Black/African and Hispanic/Latino Populations

**DOI:** 10.64898/2026.09.04.26362286

**Authors:** Lei Wang, Yanbo Zhang, Sang M Nguyen, Brandilyn A. Peters, Qibin Qi, Robert D. Burk, Robert Kaplan, Bharat Thyagarajan, Martha Daviglus, Xiao-Ou Shu, Siyuan Ma, Qi Dai, Martha J. Shrubsole, Wei Zheng, Danxia Yu

**Affiliations:** Division of Epidemiology, Department of Medicine, Vanderbilt University Medical Center, Nashville, TN, USA; Department of Epidemiology and Population Health, Albert Einstein College of Medicine, Bronx, NY, USA; Departments of Pediatrics, Microbiology & Immunology, and Obstetrics, Gynecology & Women’s Health, Albert Einstein College of Medicine, Bronx, NY, USA; Division of Public Health Sciences, Fred Hutchinson Cancer Research Center, Seattle, WA, USA; Department of Laboratory Medicine and Pathology, University of Minnesota, Minneapolis, MN, USA; Institute for Minority Health Research, University of Illinois Chicago, Chicago, IL, USA; Division of Biostatistics, Department of Medicine, Vanderbilt University Medical Center, Nashville, TN, USA; International Epidemiology Field Station, Vanderbilt University Medical Center, Nashville, TN, USA

**Keywords:** Dietary pattern, Healthy Eating Index, Dietary Approaches to Stop Hypertension, Empirical Dietary Inflammatory Potential, Empirical Dietary Index for Hyperinsulinemia, Ultra-processed foods, Gut microbiome, Black/African American, Hispanic/Latino

## Abstract

**Background:** Large-scale studies examining dietary patterns and gut microbiome have focused predominantly on European ancestry populations; evidence from other ancestry groups remains limited.

**Objectives:** We conducted a two-stage study to evaluate associations of multiple dietary patterns with gut microbiome diversity and composition among Black/African American adults and validate significant findings in a Hispanic/Latino population in the US.

**Methods:** Included were 514 Black participants from the Southern Community Cohort Study (SCCS) and 2,133 participants from the Hispanic Community Health Study/Study of Latinos (HCHS/SOL). Diet was collected by food frequency questionnaires or 24-h dietary recalls at cohort enrollment. Gut microbiome profiling was performed by shotgun metagenomic sequencing of stool samples collected during cohort follow-up. Five dietary patterns - Healthy Eating Index (HEI), Dietary Approaches to Stop Hypertension (DASH), Empirical Dietary Inflammatory Potential (EDIP), Empirical Dietary Index for Hyperinsulinemia (EDIH), and Ultra-Processed Foods (UPF) - were examined for associations with microbiome diversity and composition by linear regression after data transformation and confounders adjustment. Microbial taxa with FDR<0.1 and their constituent lower-level features identified in SCCS were targeted for validation in HCHS/SOL.

**Results:** In SCCS, HEI, DASH, EDIP, or EDIH were associated with the relative abundances of 14 microbial taxa, primarily members of families *Coriobacteriaceae* and *unclassified Firmicutes,* as well as species *Lactococcus lactis* and *Clostridium sp. AF20-17LB*. Among these, the inverse associations of genus *Collinsella* and its species *C. aerofaciens* with HEI or DASH were validated in HCHS/SOL (all *P*<0.05). Additionally, *Collinsella* and *C. aerofaciens* were associated with higher odds of obesity in SCCS (BMI≥ 30 kg/m2; OR [95%CI]:1.24 [1.01, 1.53] and 1.32 [1.07, 1.63], respectively).

**Conclusions:** Healthier dietary patterns were consistently associated with lower abundances of *Collinsella* and *C. aerofaciens* in Black/African and Hispanic/Latino Americans. Further research should clarify causal pathway linking diet, gut microbiome, and health outcomes across diverse populations.

## Introduction

Diet is a key determinant of human health and plays a central role in the development of obesity and related cardiometabolic diseases (CMD). Poor diet, such as high intake of sodium, sugar-sweetened beverages, and processed meat and low intake of whole grains, fruit, vegetables, and nuts, has been consistently linked with increased CMD risk and mortality (1). Increasing evidence suggests that modulation of the gut microbiota represents an important biological pathway linking diet to health outcomes (2).

Numerous studies have examined associations between nutrients or food groups and gut microbiome and have identified diet-related gut bacteria with beneficial or detrimental effects on human health. For example, plant-based protein and fiber intake were associated with increases in beneficial genera, including *Bifidobacterium* and *Lactobacillus*, along with higher production of anti-inflammatory short-chain fatty acids (SCFA); in contrast, diets rich in animal-based protein or saturated fats were linked to the enrichment of bile-tolerant bacteria, such as *Bacteroides*, *Alistipes*, and *Bilophila*, which may promote metabolic inflammation (3,4). Individual food groups, including red meat/processed meat, vegetables, fruits, nuts, legumes, tea, and coffee, have also been shown to influence microbial composition and function (5,6).

Because foods and nutrients are consumed in combination, their synergistic and interactive effects on the gut microbiome may be complex. Dietary pattern analysis provides a more comprehensive representation of habitual intake and may better capture diet-microbiome relationships. Studies of healthy dietary patterns, such as the Healthy Eating Index (HEI) and Dietary Approaches to Stop Hypertension (DASH), have reported higher abundances of fiber-fermenting bacteria (e.g., genera *Coprococcus* and *Lachnospira* and species *Eubacterium eligens*), primarily from the order *Clostridiales* within the phylum *Firmicutes* (7–10). These bacteria were also associated with more favorable cardiometabolic profiles, including lower levels of triglyceride, fasting insulin, waist circumference, and waist-to-hip ratio (8). To date, most large-scale, population-based studies on dietary patterns and gut microbiome have been conducted predominantly in populations of European ancestry (6,7,9–13). Relevant evidence from other racial/ethnic groups remains limited, particularly the evidence in Black/African American adults is lacking. Furthermore, studies examining the pro-inflammatory or hyperinsulinemia dietary patterns, such as the Empirical Dietary Inflammatory Potential (EDIP) and Empirical Dietary Index for Hyperinsulinemia (EDIH), as well as ultra-processed food (UPF) intake in relation to the gut microbiome are scarce and have yielded inconsistent findings (14–19). Additional studies are needed to robustly identify and validate microbiome features associated with healthy and unhealthy dietary patterns in diverse populations, particularly those underrepresented in microbiome research.

To address these gaps, we examined associations of five dietary patterns —HEI, DASH, EDIP, EDIH, and UPF intake—with gut microbiome diversity and composition among Black/African American participants from the Southern Community Cohort Study (SCCS) and sought to validate significant findings in an independent US population from the Hispanic Community Health Study/Study of Latinos (HCHS/SOL).

## Methods

### Study population

This discovery stage was conducted in the SCCS, a prospective cohort study primarily comprising Black/African American and low-income Americans (**Figure 1**) (20). In brief, 84,735 Americans aged 40-79 years old were enrolled from 12 southeastern states during March 2002 – September 2009. Structured questionnaires were administered to collect sociodemographics, diet, lifestyle factors, anthropometrics, and medical history at baseline. For the present study, we focused on 600 participants who self-reported as Black/African American, provided stool samples during the 4^th^ follow-up (2018–2021), and had these samples analyzed using shotgun metagenomic sequencing. Participants who failed to pass quality control (N=6), left >10 items blank in the food frequency questionnaire (FFQ; N=42), had inflammatory bowel disease (N=2), underwent procedures requiring bowel preparation in the two months prior to stool collection (N=30), or reported antibiotic use or diarrhea in the two months prior to stool collection (N=6) were excluded, leaving 514 participants in the final analysis.

**Figure 1.**
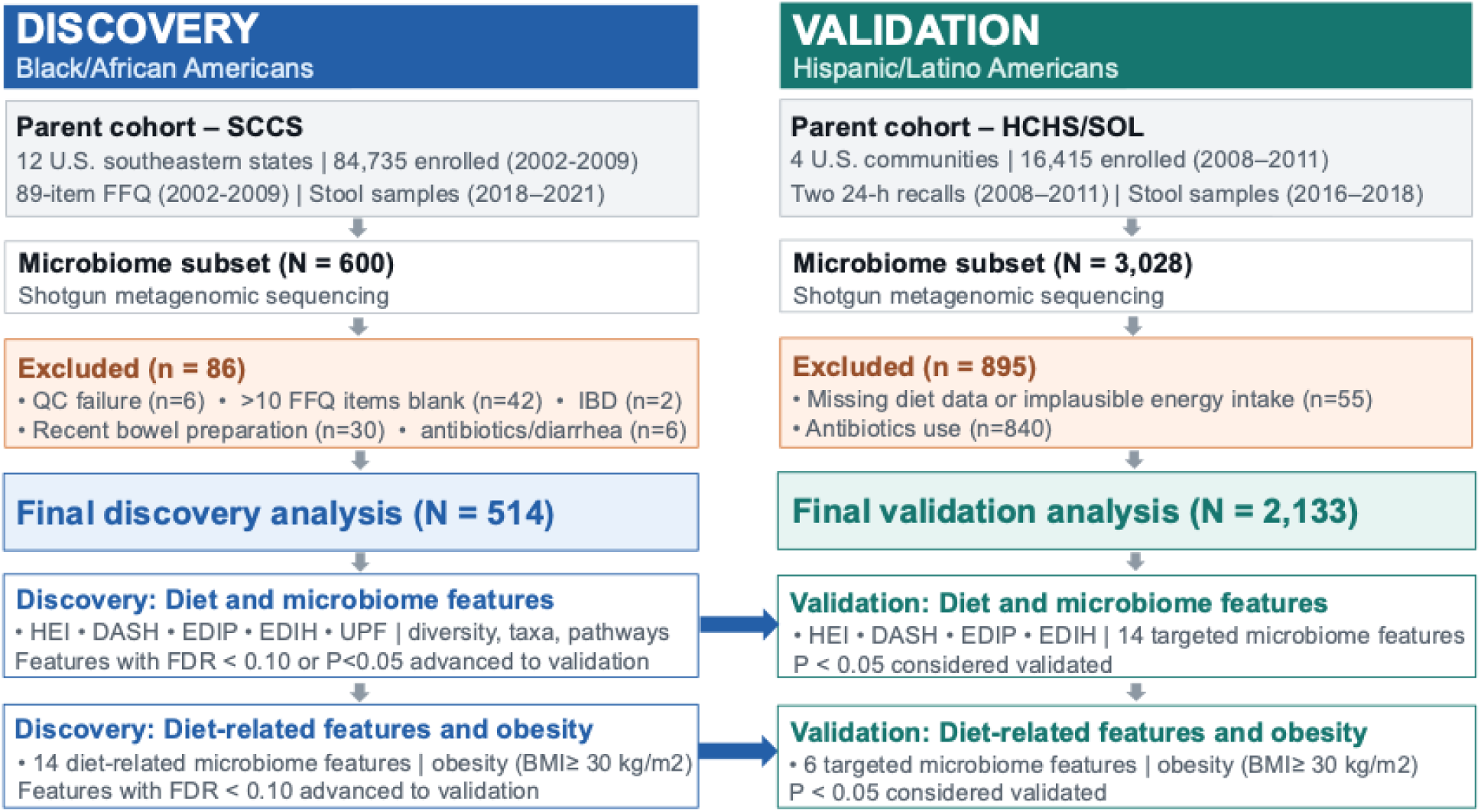
Study flowchart Abbreviations: DASH, Dietary Approaches to Stop Hypertension; EDIH, Empirical Dietary Index for Hyperinsulinemia; EDIP, Empirical Dietary Inflammatory Potential; FDR, false discovery rate; FFQ, food frequency questionnaire; HCHS/SOL, Hispanic Community Health Study / Study of Latinos; HEI, Healthy Eating Index; SCCS, Southern Community Cohort Study; UPF, ultra-processed foods.

The validation stage was conducted in the HCHS/SOL, a prospective, Hispanic/Latino population-based cohort study (21). A total of 16,415 participants aged 18-74 years were enrolled during 2008-2011 from 4 US communities (Bronx, NY; Chicago, IL; San Diego, CA; and Miami, FL). Comprehensive information, including sociodemographics, lifestyles, and medical conditions, was collected during the first (2008–2011) and second (2014–2017) clinic visits. The current validation analysis focused on 3,028 participants with gut microbiome profiling data from stool samples collected between 2016-2018. We further excluded participants with missing dietary data or implausible daily caloric intake (<600 or >8,000 kcal; N=55) and those who reported antibiotic use (N=840), leaving 2,133 participants in the final analysis.

The SCCS and HCHS/SOL had institutional review board (IRB) approval from all participating institutions, and the present study was approved by the IRB of Vanderbilt University Medical Center. All participants provided written informed consent.

### Dietary patterns

In the SCCS, dietary intake was assessed using an 89-item food frequency questionnaire (FFQ) at baseline (22). Participants reported their usual intake over the past 12 months, with consumption frequency recorded in nine categories ranging from “never” to “≥2 times per day”. Daily intake (grams/day) for each food was calculated by multiplying the reported frequency by the race-and sex-specific portion sizes (23,24). The validity of the FFQ was evaluated against 24-hour dietary recalls, with kappa coefficients ranging from 0.82 to 0.96 for macronutrients and from 0.73 to 0.95 for micronutrients (23,25). Five dietary patterns were derived based on the 89-item FFQ data, including two healthy dietary patterns (HEI and DASH) and three unhealthy dietary patterns (EDIP, EDIH, and UPF).

#### Healthy Eating Index (HEI)

The SCCS FFQ data were linked with the MyPyramid Equivalents Database from the U.S. Department of Agriculture (USDA) to generate the equivalent intake (cup or ounce equivalents per 1000 kcal) for food groups listed in the 2010 Dietary Guidelines for Americans (DGA) (26). The equivalent intake for each food group was scored according to HEI-2010 standards, which comprised 12 components with a total score ranging from 0 to 100 points, with higher scores indicating a healthier diet.(27) The HEI-2010 included nine adequacy components of total fruit (5 points), whole fruit (5 points), total vegetables (5 points), greens and beans (5 points), whole grains (10 points), dairy (10 points), total protein foods (5 points), seafood and plant proteins (5 points), and fatty acids (10 points) and three moderation components of refined grains (10 points), sodium (10 points), and calories from solid fats, alcohol, and added sugars (20 points) (26).

#### Dietary Approaches to Stop Hypertension (DASH)

The generated equivalent intake of food groups per 1000 kcal were scored for 11 components of DASH score based on the recommendations from the National Heart, Lung and Blood Institute for lowering blood pressure with DASH eating plan in 2006, including fruits (10 points), vegetables (10 points), total grains (5 points), high-fiber grains (5 points), dairy (5 points), low-fat dairy (5 points), nuts, seeds and legumes (10 points), lean meats, poultry, fish and eggs (10 points), sweets and added sugar (10 points), fats and oils (10 points), and sodium (10 points) (28,29). The total score ranged from 0 to 90 points, with higher scores indicating a healthier diet with greater alignment with the DASH diet.

#### Empirical Dietary Inflammatory Potential (EDIP)

The EDIP score was developed based on 18 food groups (9 are proinflammatory and 9 are anti-inflammatory) most predictive of 3 plasma inflammatory markers (interleukin-6, C-reactive protein, and tumor necrosis factor α receptor 2) in the Nurses’ Health Study (30). We calculated EDIP score as described in Tabung *et al*’s paper (30). In brief, the intake amounts (grams/day) of food items were converted to servings/day and grouped into 18 components (**Supplementary Table 1**). The total number of servings for each component was calculated and multiplied by the weights provided in the paper. The 18 components were summed to obtain the EDIP score first, and the score was then divided by 1000 to reduce its magnitude. A higher EDIP score indicates a less healthy diet.

#### Empirical Dietary Index for Hyperinsulinemia (EDIH)

The EDIH score was developed based on 18 food groups most predictive of fasting plasma C-peptide (13 were positively associated and 5 were inversely associated) in the Nurses’ Health Study (31). In SCCS, the intake amounts of food items were converted to servings per 1000 kcal and grouped into 18 components (**Supplementary Table 1**). The total number of servings for each component was calculated, multiplied by the weights reported in Tabung *et al*’s paper, and then summed to obtain the EDIH score (31). A higher EDIH score indicates a less healthy diet.

#### Ultra-processed foods (UPF)

Based on the Nova classification system, a validated framework for categorizing foods according to the extent and purpose of industrial processing, all FFQ items were classified into four groups (un/minimally processed, processed culinary ingredient, processed foods, and ultra-processed foods) (32). We classified 46 food items as UPF; 32 were assigned 100% weight, and 14 were assigned 20 – 70% correction weights to account for the probability that the food or food group would be ultra-processed in the SCCS population (25). UPF intake was calculated as the percentage of total daily food intake by weight (grams). Higher UPF intake indicates a less healthy diet.

In HCHS/SOL, dietary intake was assessed using two 24-hour dietary recalls at baseline. Food and nutrient intakes were analyzed using the multiple-pass methods of the Nutrition Data System for Research software version 2011 (Nutrition Coordinating Center, University of Minnesota). The average food intake from both recalls was used to calculate dietary pattern scores following the same methods as in the SCCS (**Supplementary Table 1**).

### Gut Microbiome Profiling

Stool sample collection, DNA extraction, and microbiome sequencing in the SCCS and HCHS/SOL have been described previously (8,33). In brief, in the SCCS, stool samples were collected by fecal occult blood test (FOBT) cards following a standard protocol (34). DNA was extracted using the QIAGEN DNeasy PowerSoil kit. Whole-genome shotgun metagenomic sequencing libraries were constructed using the DNBSEQ short-read library preparation kit (MGI Tech, Shenzhen, China). Sequencing was performed on the MGISEQ-2000 platform (150-bp paired-end) at BGI Americas (Cambridge, MA, USA). Taxonomic profiling from phylum to species was conducted using MetaPhlAn4 (mpa_vJun23_CHOCOPhlAnSGB_202403) (35). Functional profiling was performed using HUMAnN 3.0 (36), with UniRef90 comprehensive protein database as reference (37). Relative abundances of gut microbial metabolic pathways were estimated with the MetaCyc database as reference (38). The medium sequencing depth of included SCCS participants was 39.9M reads (IQR: 39.8-40.0M). The presence of individual microbial taxa or pathways was defined as relative abundance ≥0.001% in a sample. Taxa or pathways with a prevalence of <20% were excluded from the analysis. In HCHS/SOL, participants collected stool samples at home using stool collection kits during the second clinic visit (8). Shotgun metagenomic sequencing was performed in the Knight laboratory at the University of California, San Diego. In brief, DNA was extracted following the Earth Microbiome Project protocol. Sequencing adapters and barcode indices were added according to the iTru adapter protocol, and the amplified, indexed libraries were purified, quantified, and normalized for sequencing on the Illumina NovaSeq platform. FASTQ sequence reads were demultiplexed, adapters were trimmed, and reads mapping to the human genome were filtered out using Bowtie2. Taxonomic profiling was performed using MetaPhlAn3 (mpa_v30_CHOCOPhlAn_201901). The median sequencing depth was 795K reads (IQR: 620-973K).

### Statistical analysis

In the discovery stage (SCCS), participants’ characteristics were summarized as mean (SD) or number (percentage) and compared across tertiles of HEI or UPF score using ANOVA or chi-square test. Gut microbiome α-diversity was assessed by Shannon index and β-diversity by Bray-Curtis distance. Associations of the five dietary patterns with α-diversity were evaluated using linear regression and with β-diversity using permutational multivariate analysis of variance. Relative abundances of microbial taxa and metabolic pathways were transformed using 2-fold arcsine square root transformation [2∗asin(sqrt(x))] and standardized by mean and SD. Associations of dietary pattern scores (per 1 SD increase) with taxa and pathway abundances were analyzed using linear regression. Covariates in the models included time interval between enrollment and stool collection, enrollment source (community health center, general population), enrollment age, sex, education (less than high school, graduated high school, some college, graduated college or higher), annual household income (<$15,000, $15,000–24,999, $25,000–49,999, ≥ $50,000), smoking status (never, former, current), smoking pack years, alcohol drinking (drinks/day), physical activity (metabolic equivalent hours/day), sitting hours, total daily calories, BMI, and disease history including cancer, CVD, diabetes, hypertension, and hypercholesterolemia at baseline. We further evaluated associations of dietary pattern-related taxa with obesity status (BMI≥ 30 kg/m^2^) at the 4^th^ follow-up using logistic regression with adjustment for covariates measured near the time of stool collection. To investigate potential effect modification on diet-microbiome associations, stratification analysis was conducted by enrollment age (<53 vs. ≥53 y), sex (male vs. female), smoking status (ever vs. never), alcohol drinking (yes vs. no), physical activity (low vs. high), obesity (BMI<30 vs. ≥30 kg/m^2^), and history of diabetes, hypertension, and hypercholesterolemia. Benjamini and Hochberg false discovery rates (FDR) were calculated at each taxonomic level, and an FDR <0.10 was considered significant.

Taxa with FDR<0.1 and their constituent lower-level features identified in the discovery stage were targeted for validation in HCHS/SOL. The same data transformation (2-fold arcsine square root transformation and standardization) was applied to microbial relative abundances. Associations between dietary pattern scores and targeted taxa were evaluated by linear regression after adjusting for baseline covariates, including age, sex, study field (Bronx, Chicago, Miami, San Diego), Hispanic/Latino background (Dominican, Central American, Cuban, Mexican, Puerto Rican, South American, more than one heritage), educational attainment (less than high school, high school or equivalent, more than high school), annual household income (<$30,000, ≥$30,000), smoking status (never, former, current light, current high), alcohol drinking (never, former, current moderate, current heavy), total physical activity (metabolic equivalent hours/day), sitting hours, daily caloric intake, BMI, and history of diabetes, hypertension, hypercholesterolemia, CVD, and cancer. Associations between targeted taxa and prevalent obesity were examined by logistic regression after adjusting for the same covariates collected at the second clinic visit. A two-sided *P* value < 0.05 with same direction of association as the discovery stage was considered validated.

## Results

### Participants’ characteristics

SCCS participants had a mean (SD) age of 53.2 (7.7) y at enrollment and 66.5 (7.8) y at stool collection; the average time interval from enrollment to stool collection was 13.3 (2.0) y (**Table 1**). Participants with a healthier diet (i.e., higher HEI score/lower UPF intake) were more likely to be older, female, non-smokers, and non-drinkers and have higher education and income levels and lower daily energy intake compared to those with an unhealthier diet (*P*<0.05). We also found that participants with a healthier diet had a higher prevalence of cancer or hypercholesterolemia, indicating a possible dietary change after disease diagnosis (**Table 1**). The HEI showed a strong positive correlation with DASH (r=0.82) and moderate-to-strong negative correlations with EDIP (r=-0.34), EDIH (r=-0.57), and UPF (r=-0.66; **Supplementary Figure 1**).

**Table 1.** Baseline characteristics of study participants across HEI or UPF tertiles in the SCCS

| Baseline characteristics | All<br>(N=514) | HEI |  |  |  | UPF <sup>1</sup> |  |  |  |
| --- | --- | --- | --- | --- | --- | --- | --- | --- | --- |
|  |  | Tertile 1<br>(22-55 points)<br>*Unhealthiest* | Tertile 2<br>(55-67 point) | Tertile 3<br>(67-92 points)<br>*Healthiest* | *P* value | Tertile 1<br>(2.2-31.2%)<br>*Healthiest* | Tertile 2<br>(31.3-44.9%) | Tertile 3<br>(45.0-83.6%)<br>*Unhealthiest* | *P* value |
| **Enrollment age, years, mean (SD)** | 53.2 (7.7) | 50.7 (6.5) | 52.5 (7.3) | 56.4 (8.1) | **<.0001** | 55.1 (8.1) | 53.4 (7.5) | 51.1 (6.8) | **<.0001** |
| **Age at stool collection, years, mean (SD)** | 66.5 (7.8) | 64.0 (6.6) | 65.9 (7.5) | 69.6 (8.1) | **<.0001** | 68.2 (8.2) | 66.7 (7.9) | 64.6 (6.8) | **<.0001** |
| **Time interval from enrollment to stool collection, years, mean (SD)** | 13.3 (2.0) | 13.2 (2.2) | 13.4 (2.0) | 13.2 (1.7) | 0.47 | 13.0 (1.8) | 13.3 (2.0) | 13.5 (2.0) | 0.13 |
| **Sex, n (%)** |  |  |  |  | **0.0001** |  |  |  | **0.02** |
| Male | 144 (28.0) | 65 (38.0) | 42 (24.6) | 37 (21.5) |  | 36 (21.1) | 49 (28.7) | 59 (34.3) |  |
| Female | 370 (72.0) | 106 (62.0) | 129 (75.4) | 135 (78.5) |  | 135 (78.9) | 122 (71.3) | 113 (65.7) |  |
| **CHC enrollment, n (%)** | 400 (77.8) | 137 (80.1) | 143 (83.6) | 120 (70.0) | **0.006** | 128 (74.9) | 129 (75.4) | 143 (83.1) | 0.12 |
| **Educational attainment, n (%)** |  |  |  |  | **0.006** |  |  |  | **0.03** |
| <12 years | 83 (16.2) | 25 (14.6) | 37 (21.6) | 21 (12.2) |  | 27 (15.8) | 24 (14.0) | 32 (18.6) |  |
| Graduated high school | 178 (34.6) | 74 (43.3) | 52 (30.4) | 52 (30.2) |  | 51 (29.8) | 64 (37.4) | 63 (36.6) |  |
| Some college | 164 (31.9) | 56 (32.7) | 53 (31.0) | 55 (32.0) |  | 53 (31.0) | 50 (29.2) | 61 (35.5) |  |
| Graduated college or higher | 89 (17.3) | 16 (9.4) | 29 (17.0) | 44 (25.6) |  | 40 (23.4) | 33 (19.3) | 16 (9.3) |  |
| **Annual household income, n (%)** |  |  |  |  | **0.003** |  |  |  | 0.43 |
| <\$15,000 | 217 (42.2) | 84 (49.1) | 77 (45.0) | 56 (32.6) | | 67 (39.2) | 67 (39.2) | 83 (48.3) | |
| \$15,000 - \$24,999 | 129 (25.1) | 42 (24.6) | 46 (26.9) | 41 (23.8) | | 42 (24.5) | 45 (26.3) | 42 (24.4) | |
| \$25,000 - \$49,999 | 97 (18.9) | 31 (18.1) | 22 (12.9) | 44 (25.6) | | 33 (19.3) | 34 (19.9) | 30 (17.4) | |
| ≥ \$50,000 | 71 (13.8) | 14 (8.2) | 26 (15.2) | 31 (18.0) | | 29 (17.0) | 25 (14.6) | 17 (9.9) | |
| **Deprivation index, mean (SD)** | 0.80 (1.14) | 0.94 (1.1) | 0.87 (1.2) | 0.61 (1.1) | **0.02** | 0.59 (1.08) | 0.91 (1.17) | 0.91 (1.14) | **0.01** |
| **Smoking status, n (%)** |  |  |  |  | **<.0001** |  |  |  | **0.006** |
| Never | 243 (47.3) | 65 (38.0) | 79 (46.2) | 99 (57.6) |  | 94 (55.0) | 74 (43.3) | 75 (43.6) |  |
| Former | 133 (25.8) | 40 (23.4) | 41 (24.0) | 52 (30.2) |  | 46 (26.9) | 50 (29.2) | 37 (21.5) |  |
| Current | 138 (26.9) | 66 (38.6) | 51 (29.8) | 21 (12.2) |  | 31 (18.1) | 47 (27.5) | 60 (34.9) |  |
| **Smoking pack-years among ever-smokers, mean (SD)** | 17.3 (16.6) | 19.7 (18.3) | 16.5 (14.9) | 15.0 (16.0) | 0.16 | 15.9 (13.7) | 18.0 (17.1) | 17.8 (18.2) | 0.65 |
| **Alcohol drinking <sup>2</sup>, n (%)** |  |  |  |  | **0.0002** |  |  |  | 0.19 |
| None | 259 (50.4) | 76 (44.4) | 82 (48.0) | 101 (58.7) |  | 94 (55.0) | 83 (48.5) | 82 (47.7) |  |
| Moderate | 185 (36.0) | 64 (37.4) | 57 (33.3) | 64 (37.2) |  | 61 (35.7) | 65 (38.0) | 59 (34.3) |  |
| Heavy | 70 (13.6) | 31 (18.1) | 32 (18.7) | 7 (4.1) |  | 16 (9.4) | 23 (13.5) | 31 (18.0) |  |
| **Number of drinks/day among alcohol drinkers, mean (SD)** | 1.9 (4.4) | 3.0 (6.1) | 1.8 (3.4) | 0.6 (1.2) | **0.002** | 1.4 (4.0) | 1.4 (3.1) | 2.8 (5.5) | **0.04** |
| **Total physical activity, MET-hours/day, mean (SD)** | 21.7 (17.0) | 22.9 (17.4) | 22.3 (17.9) | 20.1 (15.5) | 0.28 | 20.6 (16.1) | 21.7 (16.7) | 22.9 (18.1) | 0.46 |
| **Total sitting time, hours/day, mean (SD)** | 9.6 (4.4) | 9.9 (4.5) | 9.8 (4.4) | 9.0 (4.3) | 0.18 | 9.0 (4.4) | 9.8 (4.4) | 9.8 (4.6) | 0.17 |
| **Total energy intake, kcal/day, mean (SD)** | 2437 (1358) | 2530 (1306) | 2596 (1511) | 2188 (1212) | **0.01** | 2180 (1231) | 2487 (1451) | 2644 (1349) | **0.006** |
| **BMI, kg/m<sup>2</sup>, mean (SD)** | 30.9 (6.8) | 30.4 (7.3) | 31.9 (7.1) | 30.5 (5.9) | 0.05 | 30.7 (6.6) | 30.9 (6.8) | 31.1 (7.0) | 0.84 |
| **Prevalent medical conditions, n (%)** |  |  |  |  |  |  |  |  |  |
| Cancer | 34 (6.6) | 13 (7.6) | 5 (2.9) | 16 (9.3) | **0.049** | 18 (10.5) | 7 (4.1) | 9 (5.2) | **0.04** |
| CVD (CHD or stroke) | 36 (7.0) | 9 (5.3) | 12 (7.0) | 15 (8.7) | 0.46 | 15 (8.8) | 11 (6.4) | 10 (5.8) | 0.53 |
| Diabetes | 80 (15.6) | 21 (12.3) | 26 (15.2) | 33 (19.2) | 0.21 | 26 (15.2) | 24 (14.0) | 30 (17.4) | 0.68 |
| Hypertension | 286 (55.6) | 86 (50.3) | 101 (59.1) | 99 (57.6) | 0.22 | 98 (57.3) | 94 (55.0) | 94 (54.7) | 0.86 |
| Hypercholesterolemia | 200 (38.9) | 46 (26.9) | 64 (37.4) | 90 (52.3) | **<.0001** | 74 (43.3) | 77 (45.0) | 49 (28.5) | **0.003** |

Abbreviations: BMI, body mass index; CHC, Community Health Center; HEI, Healthy Eating Index; MET, metabolic equivalent of task; SCCS, Southern Community Cohort Study; SD, standard deviation; UPF, ultra-processed foods.
<sup>1</sup>Intake of UPF was estimated as the proportion of UPF in daily food intake (% grams).
<sup>2</sup>Heavy drinking was defined as alcohol consumption of >2 drinks per day in men or alcohol consumption of >1 drink per day in women; moderate drinking was defined as alcohol consumption of >0 to ≤2 drinks per day in men or >0 to ≤1 drink per day in women.

HCHS/SOL participants had a mean (SD) age of 50.8 (11.1) y at enrollment and 58.1 (11.2) y at stool collection. Compared to the SCCS participants, HCHS/SOL participants had lower education levels, alcohol use, sitting time, and daily calories intake, but higher smoking rate and physical activity levels (**Supplementary Table 2**). The mean HEI score was similar between two cohorts, whereas SCCS had a higher DASH score and lower EDIP and EDIH scores than HCHS/SOL.

### Dietary patterns and gut microbiome diversity and composition

In the SCCS, none of the five dietary patterns was significantly associated with the Shannon index or Bray–Curtis distance (variation explained: 0.18%-0.26%) after adjusting for covariates (all *P* values >0.05; **Supplementary Figure 2 & 3**). Age, sex, physical activity, and history of diabetes were significantly associated with the Bray–Curtis distance, with these factors explaining 0.28% to 0.52% of the variation (all *P* values < 0.05).

We evaluated associations of the five dietary patterns with 10 phyla, 65 classes, 71 orders, 92 families, 212 genera, and 338 species in the SCCS. Higher HEI or DASH score was associated with decreased abundances of families *Coriobacteriaceae* (β=-0.165 for HEI and -0.160 for DASH) and *unclassified Firmicutes* (β=-0.152 for HEI and -0.169 for DASH), while higher EDIP was associated with increased abundances of family *Coriobacteriaceae* (β=0.164) and genus *GGB38744* (β=0.163, all *FDR*<0.10; **Table 2**). Higher EDIP also showed significant associations with decreased abundances of genus *Lactococcus* (β=-0.210) and its species *lactis* (β=-0.218), and higher EDIH was associated with a decreased abundance of species *Clostridium sp. AF20-17LB* (β=-0.214). No significant associations were found for the UPF and gut microbial taxa. We further explored which taxa within families *Coriobacteriaceae, unclassified Firmicutes,* or genus *GGB38744* were driving the associations with HEI, DASH, or EDIP. We found inverse associations of genus *Collinsella* and its species *aerofaciens* with HEI and DASH, and positive associations with EDIP (all *P*<0.05, *FDR*>0.1; **Table 2**). Additionally, genus *GGB9512* and its species *SGB14909* within family *unclassified Firmicutes* were negatively associated with DASH, and species *SGB14842* within genus *GGB38744* was positively associated with EDIP (all *P*<0.05, *FDR*>0.1; **Table 2**). A total of 427 microbial metabolic pathways were included in the SCCS analysis, but no significant association was found with any of the dietary patterns (all *FDR*>0.1).

**Table 2.**
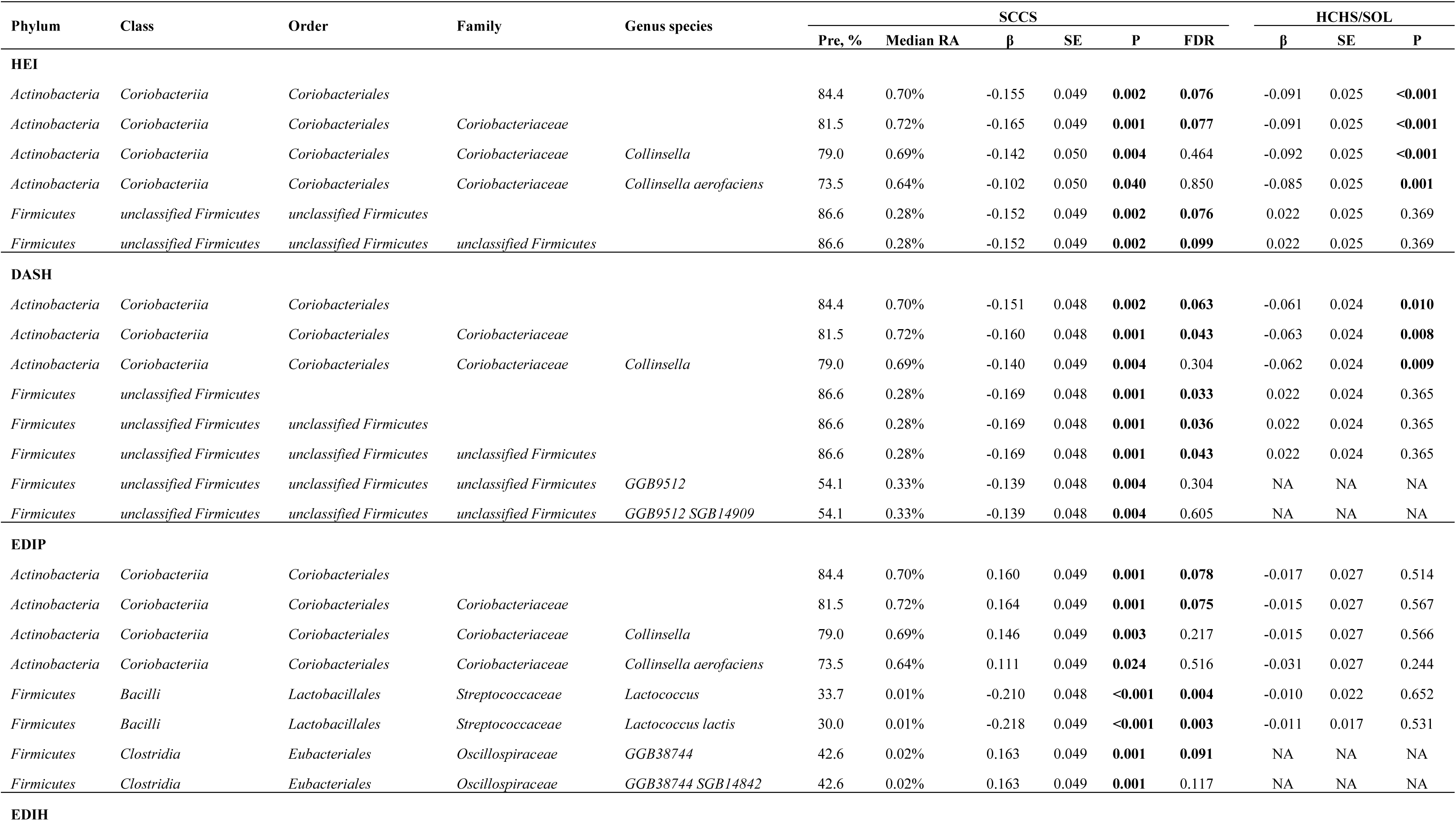
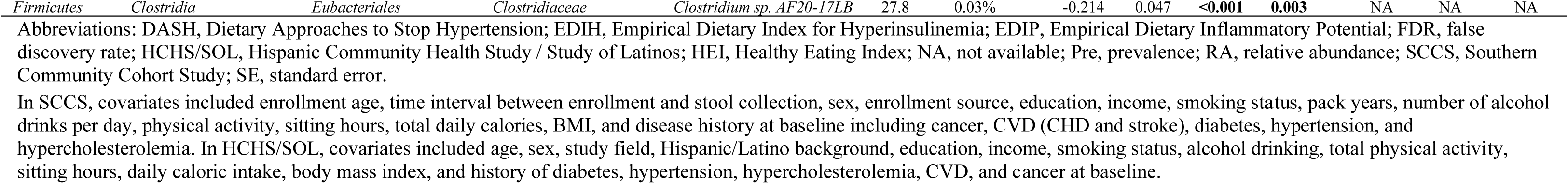
Discovery and validation of gut microbial taxa related to dietary patterns

| Phylum | Class | Order | Family | Genus species | SCCS |  |  |  |  | HCHS/SOL |  |  |  |
| --- | --- | --- | --- | --- | --- | --- | --- | --- | --- | --- | --- | --- | --- |
|  |  |  |  |  | Pre, % | Median RA | β | SE | P | FDR | β | SE | P |
| HEI |  |  |  |  |  |  |  |  |  |  |  |  |  |
| Actinobacteria | Coriobacteriia | Coriobacteriales |  |  | 84.4 | 0.70% | -0.155 | 0.049 | 0.002 | 0.076 | -0.091 | 0.025 | <0.001 |
| Actinobacteria | Coriobacteriia | Coriobacteriales | Coriobacteriaceae |  | 81.5 | 0.72% | -0.165 | 0.049 | 0.001 | 0.077 | -0.091 | 0.025 | <0.001 |
| Actinobacteria | Coriobacteriia | Coriobacteriales | Coriobacteriaceae | Collinsella | 79.0 | 0.69% | -0.142 | 0.050 | 0.004 | 0.464 | -0.092 | 0.025 | <0.001 |
| Actinobacteria | Coriobacteriia | Coriobacteriales | Coriobacteriaceae | Collinsella aerofaciens | 73.5 | 0.64% | -0.102 | 0.050 | 0.040 | 0.850 | -0.085 | 0.025 | 0.001 |
| Firmicutes | unclassified Firmicutes | unclassified Firmicutes |  |  | 86.6 | 0.28% | -0.152 | 0.049 | 0.002 | 0.076 | 0.022 | 0.025 | 0.369 |
| Firmicutes | unclassified Firmicutes | unclassified Firmicutes | unclassified Firmicutes |  | 86.6 | 0.28% | -0.152 | 0.049 | 0.002 | 0.099 | 0.022 | 0.025 | 0.369 |
| DASH |  |  |  |  |  |  |  |  |  |  |  |  |  |
| Actinobacteria | Coriobacteriia | Coriobacteriales |  |  | 84.4 | 0.70% | -0.151 | 0.048 | 0.002 | 0.063 | -0.061 | 0.024 | 0.010 |
| Actinobacteria | Coriobacteriia | Coriobacteriales | Coriobacteriaceae |  | 81.5 | 0.72% | -0.160 | 0.048 | 0.001 | 0.043 | -0.063 | 0.024 | 0.008 |
| Actinobacteria | Coriobacteriia | Coriobacteriales | Coriobacteriaceae | Collinsella | 79.0 | 0.69% | -0.140 | 0.049 | 0.004 | 0.304 | -0.062 | 0.024 | 0.009 |
| Firmicutes | unclassified Firmicutes |  |  |  | 86.6 | 0.28% | -0.169 | 0.048 | 0.001 | 0.033 | 0.022 | 0.024 | 0.365 |
| Firmicutes | unclassified Firmicutes | unclassified Firmicutes |  |  | 86.6 | 0.28% | -0.169 | 0.048 | 0.001 | 0.036 | 0.022 | 0.024 | 0.365 |
| Firmicutes | unclassified Firmicutes | unclassified Firmicutes | unclassified Firmicutes |  | 86.6 | 0.28% | -0.169 | 0.048 | 0.001 | 0.043 | 0.022 | 0.024 | 0.365 |
| Firmicutes | unclassified Firmicutes | unclassified Firmicutes | unclassified Firmicutes | GGB9512 | 54.1 | 0.33% | -0.139 | 0.048 | 0.004 | 0.304 | NA | NA | NA |
| Firmicutes | unclassified Firmicutes | unclassified Firmicutes | unclassified Firmicutes | GGB9512 SGB14909 | 54.1 | 0.33% | -0.139 | 0.048 | 0.004 | 0.605 | NA | NA | NA |
| EDIP |  |  |  |  |  |  |  |  |  |  |  |  |  |
| Actinobacteria | Coriobacteriia | Coriobacteriales |  |  | 84.4 | 0.70% | 0.160 | 0.049 | 0.001 | 0.078 | -0.017 | 0.027 | 0.514 |
| Actinobacteria | Coriobacteriia | Coriobacteriales | Coriobacteriaceae |  | 81.5 | 0.72% | 0.164 | 0.049 | 0.001 | 0.075 | -0.015 | 0.027 | 0.567 |
| Actinobacteria | Coriobacteriia | Coriobacteriales | Coriobacteriaceae | Collinsella | 79.0 | 0.69% | 0.146 | 0.049 | 0.003 | 0.217 | -0.015 | 0.027 | 0.566 |
| Actinobacteria | Coriobacteriia | Coriobacteriales | Coriobacteriaceae | Collinsella aerofaciens | 73.5 | 0.64% | 0.111 | 0.049 | 0.024 | 0.516 | -0.031 | 0.027 | 0.244 |
| Firmicutes | Bacilli | Lactobacillales | Streptococcaceae | Lactococcus | 33.7 | 0.01% | -0.210 | 0.048 | <0.001 | 0.004 | -0.010 | 0.022 | 0.652 |
| Firmicutes | Bacilli | Lactobacillales | Streptococcaceae | Lactococcus lactis | 30.0 | 0.01% | -0.218 | 0.049 | <0.001 | 0.003 | -0.011 | 0.017 | 0.531 |
| Firmicutes | Clostridia | Eubacteriales | Oscillospiraceae | GGB38744 | 42.6 | 0.02% | 0.163 | 0.049 | 0.001 | 0.091 | NA | NA | NA |
| Firmicutes | Clostridia | Eubacteriales | Oscillospiraceae | GGB38744 SGB14842 | 42.6 | 0.02% | 0.163 | 0.049 | 0.001 | 0.117 | NA | NA | NA |
| **EDIH** |  |  |  |  |  |  |  |  |  |  |  |  |  |
| *Firmicutes* | *Clostridia* | *Eubacteriales* | *Clostridiaceae* | *Clostridium sp. AF20-17LB* | 27.8 | 0.03% | -0.214 | 0.047 | **<0.001** | **0.003** | NA | NA | NA |

Abbreviations: DASH, Dietary Approaches to Stop Hypertension; EDIH, Empirical Dietary Index for Hyperinsulinemia; EDIP, Empirical Dietary Inflammatory Potential; FDR, false discovery rate; HCHS/SOL, Hispanic Community Health Study / Study of Latinos; HEI, Healthy Eating Index; NA, not available; Pre, prevalence; RA, relative abundance; SCCS, Southern Community Cohort Study; SE, standard error.
In SCCS, covariates included enrollment age, time interval between enrollment and stool collection, sex, enrollment source, education, income, smoking status, pack years, number of alcohol drinks per day, physical activity, sitting hours, total daily calories, BMI, and disease history at baseline including cancer, CVD (CHD and stroke), diabetes, hypertension, and hypercholesterolemia. In HCHS/SOL, covariates included age, sex, study field, Hispanic/Latino background, education, income, smoking status, alcohol drinking, total physical activity, sitting hours, daily caloric intake, body mass index, and history of diabetes, hypertension, hypercholesterolemia, CVD, and cancer at baseline.

Taxa identified in the SCCS at FDR<0.1, along with their constituent lower-level features that met nominal significance criteria (*P*<0.05 but *FDR*>0.1), were carried forward for validation in HCHS/SOL. Among these, the inverse associations of HEI and DASH with order *Coriobacteriales* and its family *Coriobacteriaceae*, genus *Collinsella*, and species *aerofaciens* were validated (all *P* <0.05; **Table 2**).

### Dietary pattern-related bacteria and obesity

A total of 266 (52%) SCCS participants had obesity at the 4^th^ follow-up, and 910 (43%) HCHS/SOL participants had obesity at second clinic visit. We found that order *Coriobacteriales,* along with its family *Coriobacteriaceae*, genus *Collinsella*, and species *aerofaciens,* was associated with higher odds of obesity in the SCCS (OR=1.24 to 1.32, *FDR*<0.10; **Table 3**). Additionally, genus *GGB9512* and its species *SGB14909* within the order *unclassified Firmicutes* were also linked to a higher obesity risk (OR [95%CI] =1.26 [1.03, 1.55], *FDR*=0.09). However, none of these associations were validated in HCHS/SOL (**Table 3**). In the mediation analysis conducted in the SCCS, we found that species *Collinsella aerofaciens* mediated 90.7% and 43.2% of the associations of HEI and EDIP with obesity, respectively (*P*_ACEM_=0.04 for HEI and 0.01 for EDIP; **Supplementary Figure 4**).

**Table 3.**
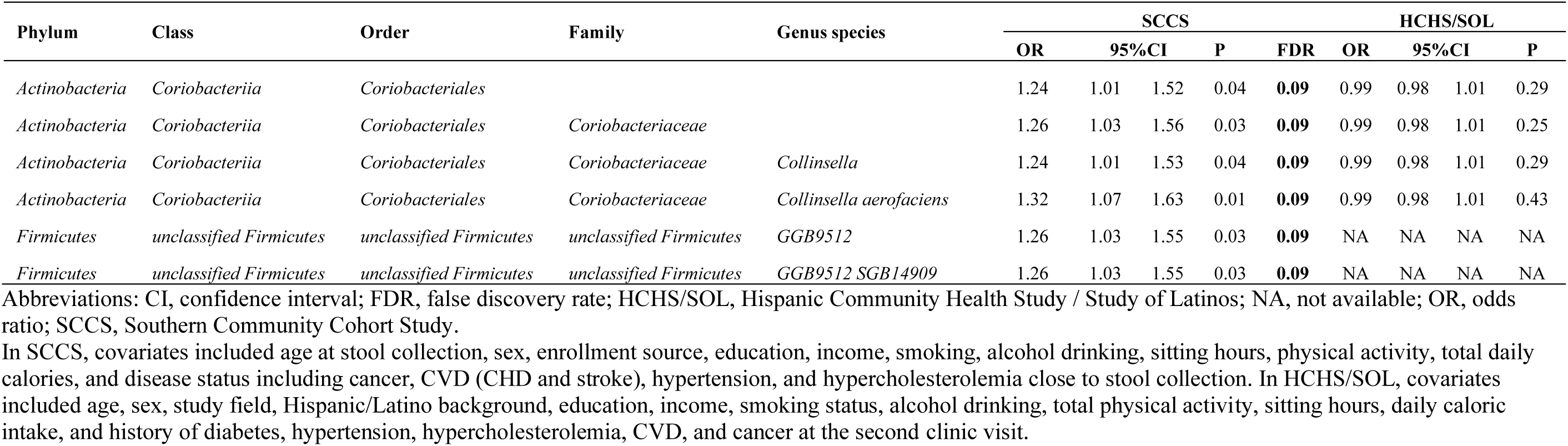
Discovery and validation of diet-related gut microbial taxa with obesity risk

### Effect modifications on diet-taxa associations in SCCS

We conducted subgroup analyses in SCCS and found potential modification effects by demographics (i.e., sex), lifestyle factors (i.e., smoking, alcohol drinking, and physical activity), and metabolic disease status (i.e., obesity and diabetes) on the diet-microbiome associations (**Table 4**). The associations of EDIP with family *Desulfovibrionaceae* and its genus *Desulfovibrio* were positive among males, while negative among females (*FDR* for interaction=0.07 and 0.09). Additionally, the associations between EDIP and several genera (i.e., *Lactococcus, Guopingia, Anaerofustis,* and *Intestinibacter*) and species (i.e., *Lactococcus lactis* and *Intestinibacter bartlettii*) were more evident among never-smokers, non-drinkers, or individuals with high physical activity compared to their counterparts (all *FDR* for interaction<0.1). DASH was associated with an increased abundance of species *Clostridium sp. AM22-11AC* from order *Eubacteriales* only among non-obese individuals. Furthermore, EDIP/EDIH was associated with decreased abundances of family *Eubacteriales incertae sedis* and several genera and species from order *Eubacteriales* only among individuals with diabetes (**Table 4**).

**Table 4.** Associations of dietary patterns with gut microbial taxa in participant subgroups in the SCCS

| Phylum | Class | Order | Family | Genus species | $\beta_1$ (se) | $P_1$ | $\beta_2$ (se) | $P_2$ | FDR<sub>int</sub> |
| --- | --- | --- | --- | --- | --- | --- | --- | --- | --- |
| ***DASH*** |  |  |  |  |  |  |  |  |  |
| **Smoking** |  |  |  |  | **Never** |  | **Ever** |  |  |
| *Firmicutes* | *Clostridia* | *Eubacteriales* | *Oscillospiraceae* | *Vescimonas* | -0.226 (0.069) | 0.001 | 0.135 (0.068) | 0.047 | 0.028 |
| *Firmicutes* | *Clostridia* | *Eubacteriales* | *Oscillospiraceae* | *Vescimonas coprocola* | -0.21 (0.066) | 0.002 | 0.158 (0.069) | 0.023 | 0.021 |
| **Obesity** |  |  |  |  | **No** |  | **Yes** |  |  |
| *Firmicutes* | *Clostridia* | *Eubacteriales* | *Clostridiaceae* | *Clostridium sp. AM22-11AC* | 0.168 (0.063) | 0.008 | -0.126 (0.074) | 0.089 | 0.090 |
| ***EDIP*** |  |  |  |  |  |  |  |  |  |
| **Sex** |  |  |  |  | **Men** |  | **Women** |  |  |
| *Proteobacteria* | *Deltaproteobacteria* | *Desulfovibrionales* | *Desulfovibrionaceae* |  | 0.187 (0.079) | 0.020 | -0.112 (0.057) | 0.050 | 0.073 |
| *Proteobacteria* | *Deltaproteobacteria* | *Desulfovibrionales* | *Desulfovibrionaceae* | *Desulfovibrio* | 0.247 (0.088) | 0.006 | -0.1 (0.055) | 0.072 | 0.093 |
| **Smoking** |  |  |  |  | **Never** |  | **Ever** |  |  |
| *Firmicutes* | *Bacilli* | *Lactobacillales* | *Streptococcaceae* | *Lactococcus* | -0.328 (0.075) | <0.001 | -0.014 (0.051) | 0.782 | 0.015 |
| *Firmicutes* | *Bacilli* | *Lactobacillales* | *Streptococcaceae* | *Lactococcus lactis* | -0.358 (0.076) | <0.001 | -0.015 (0.05) | 0.767 | 0.006 |
| *Firmicutes* | *Clostridia* | *Eubacteriales* | *Christensenellaceae* | *Guopingia* | -0.16 (0.071) | 0.024 | 0.11 (0.061) | 0.072 | 0.099 |
| *Firmicutes* | *Clostridia* | *Eubacteriales* | *Eubacteriaceae* | *Anaerofustis* | -0.23 (0.078) | 0.003 | 0.065 (0.053) | 0.217 | 0.090 |
| *Firmicutes* | *Clostridia* | *Eubacteriales* | *Peptostreptococcaceae* | *Intestinibacter* | -0.202 (0.071) | 0.005 | 0.105 (0.06) | 0.081 | 0.051 |
| *Firmicutes* | *Clostridia* | *Eubacteriales* | *Peptostreptococcaceae* | *Intestinibacter bartlettii* | -0.202 (0.071) | 0.005 | 0.105 (0.06) | 0.081 | 0.081 |
| **Alcohol** |  |  |  |  | **No** |  | **Yes** |  |  |
| *Firmicutes* | *Clostridia* | *Eubacteriales* | *Peptostreptococcaceae* | *Intestinibacter* | -0.242 (0.069) | 0.001 | 0.113 (0.062) | 0.070 | 0.099 |
| **Physical activity** |  |  |  |  | **Low** |  | **High** |  |  |
| *Firmicutes* | *Bacilli* | *Lactobacillales* | *Streptococcaceae* | *Lactococcus lactis* | 0.003 (0.054) | 0.957 | -0.297 (0.071) | <0.001 | 0.070 |
| **Diabetes** |  |  |  |  | **No** |  | **Yes** |  |  |
| *Firmicutes* | *Clostridia* | *Eubacteriales* | *Eubacteriales incertae sedis* |  | 0.052 (0.049) | 0.289 | -0.445 (0.149) | 0.004 | 0.058 |
| ***EDIH*** |  |  |  |  |  |  |  |  |  |
| **Diabetes** |  |  |  |  | **No** |  | **Yes** |  |  |
| *Firmicutes* | *Clostridia* | *Eubacteriales* | *Eubacteriales incertae sedis* |  | 0.046 (0.052) | 0.372 | -0.419 (0.117) | 0.001 | 0.007 |
| *Firmicutes* | *Clostridia* | *Eubacteriales* | *Eubacteriales incertae sedis* | *Candidatus Cibionibacter* | 0.046 (0.052) | 0.372 | -0.419 (0.117) | 0.001 | 0.017 |
| *Firmicutes* | *Clostridia* | *Eubacteriales* | *Eubacteriales incertae sedis* | *Candidatus Cibionibacter quicibialis* | 0.046 (0.052) | 0.372 | -0.419 (0.117) | 0.001 | 0.013 |
| *Firmicutes* | *Clostridia* | *Eubacteriales* | *Oscillospiraceae* | *unclassified Oscillospiraceae Clostridium methylpentosum* | 0.065 (0.041) | 0.118 | -0.476 (0.205) | 0.024 | 0.012 |
| *Firmicutes* | *Clostridia* | *Eubacteriales* | *Oscillospiraceae* | *unclassified Oscillospiraceae*<br>*Oscillospiraceae bacterium*<br>*CLA-AA-H250* | 0.077 (0.052) | 0.142 | -0.259 (0.112) | 0.024 | 0.096 |

Mutually adjusted for enrollment age, time interval between enrollment and stool collection, sex, enrollment source, education, income, smoking status, pack years, number of alcohol drinks per day, physical activity, sitting hours, total daily calories, BMI, and disease history at baseline including cancer, CVD (CHD and stroke), diabetes, hypertension, and dyslipidemia.

## Discussion

In this two-stage study involving Black/African and Hispanics/Latino Americans, we identified and validated several bacteria (i.e., order *Coriobacteriales*, family *Coriobacteriaceae*, genus *Collinsella*, and its species *C. aerofaciens*) that were inversely associated with healthy dietary patterns (i.e., HEI and DASH). In the SCCS, these bacteria were further linked to higher odds of obesity and mediated 43-91% of the association between diet and obesity. In addition, we observed that demographic factors, lifestyle behaviors, and metabolic disease history may modify diet-microbiome associations.

Although diet is recognized as one of the most important modifiable determinants influencing the gut microbiome, evidence linking dietary patterns to overall microbiome diversity remains inconsistent, and there is no consensus that greater diversity necessarily reflects a healthier microbiome. In the Multiethnic Cohort Study (US; n=1735-5936; White, Black, Native Hawaiian, Japanese American, and Latino) (7,9), the TwinsUK cohort (UK; n=2070; 99% White) (39), and the American Gut Project (US; n=432; 89% White) (10), several dietary patterns (i.e., HEI, DASH, and Mediterranean Diet) showed significant associations with alpha or beta diversity. EDIP/EDIH showed marginally significant associations with alpha diversity in the TwinsUK cohort (n=1610) (14) and the Osteoporotic Fractures in Men study (MrOS; US; n=795; 90% White) (15). However, HCHS/SOL (US; n=2444; Hispanic/Latino) and Malmö Offspring Study (Sweden; n=1726) reported null associations between diet (i.e., HEI, Mediterranean Diet, the healthful plant-based diet, or the health-conscious diet) and gut microbiome diversity (8,40). Furthermore, no significant association was found for UPF and alpha or beta diversity in the PREDIMED-Plus randomized clinical trial (Spain; n=385-656) (18,19) and the Obekit trial (Spain; n=359) (16). Consistent with these null findings, in our study among 514 Black/African American adults, none of the five dietary patterns (i.e., HEI, DASH, EDIP, EDIH, and UPF) was significantly associated with gut microbiome alpha or beta diversity.

Our study confirmed inverse associations between healthy dietary patterns and decreased abundance of genus *Collinsella* and its species *C. aerofaciens*, aligning with findings from the Multiethnic Cohort Study and HCHS/SOL (7–9). The genus *Collinsella* and its species *C. aerofaciens* were further linked to an increased risk of obesity in our discovery population, but not in the validation population, which could be due to population differences (Black/African American vs. US Hispanics/Latinos). Nevertheless, several studies have reported positive associations between *Collinsella* abundance and obesity (41–43). Additionally, *Collinsella* has been associated with increased inflammation and altered fat metabolism, potentially contributing to obesity-related metabolic disorders, such as insulin resistance, type 2 diabetes, and high blood pressure (8,44–47). Beyond depletion of *Collinsella*, several other gut microbial taxa have been linked with HEI or DASH in the literature, such as enrichment of *Anaerostipes, Coprococcus, Lachnospira, Faecalibacterium, Ruminococcus, Eubacterium eligens, Roseburia hominis, Butyrivibrio crossotus* from order *Clostridiales* of phylum *Firmicutes*, all known for fiber fermentation and SCFA production with anti-inflammatory properties (6–9). Two studies conducted in the TwinsUK cohort and MrOS identified 32 and 65 genera for EDIP and 15 and 46 genera for EDIH, respectively, using elastic net regression (14,15). Genera commonly identified in both studies included *Escherichia/Shigella*, *Negativibacillus, Ruminococcaceae_UCG-014*, and *Ruminococcaceae_UCG-008* for EDIP and *Faecalitalea, Negativibacillus, Turicibacter, UC5–1-2E3, Parasutterella, and Lactobacillus, Marvinbryantia, and Fournierella* for EDIH, none of which were replicated in our study. Furthermore, we did not find any microbial taxa significantly associated with UPF intake. Previous studies conducted in Brazil and Spain (n=59-656) have reported inconsistent findings (16–19). For example, a cross-sectional study of 359 Spanish participants found an increased abundance of genus *Parabacteroides* with higher UPF intake evaluated as servings/day, while a longitudinal analysis of 385 Spanish participants showed a significant decrease in *Parabacteroides spp.* with increased UPF intake evaluated as g/day (16,19). These mixed findings highlight the complexity of diet-microbiome interactions and suggest that factors such as study population characteristics, dietary assessment methods, and microbiome profiling techniques may affect observed associations.

Several exploratory findings emerged from the stratification analysis in the SCCS. The associations between dietary patterns and certain bacteria from phylum *Firmicutes* differed by lifestyle factors or metabolic disease status. For example, we observed negative associations of EDIP with *Lactococcus lactis* (a bacterium with probiotic potential) and *Intestinibacter* (a fiber-fermenting and SCFA-producing genus) among never-smokers, non-drinkers, or individual with high physical activity, but not among ever-smokers, alcohol drinkers, or those with low physical activity, suggesting that unhealthy lifestyles may confound the impact of a pro-inflammatory diet on the gut microbiome (48,49). In addition, we found that EDIP/EDIH was associated with decreased abundances of several unclassified taxa from family *Oscillospiraceae* and order *Eubacteriales* (fiber-fermenting and SCFA-producing bacteria) exclusively among diabetes patients. The systemic inflammation, insulin resistance, pre-existing dysbiosis, and impaired host–microbiome interactions in individuals with diabetes may amplify the microbiome’s vulnerability to the poor diet (50).

This study has several strengths. First, it is one of the first studies to identify gut microbes related to multiple dietary patterns and potential modifications by other host characteristics in a low-income Black/African American population, providing valuable insights into the diet-microbiome-disease interactions in this underserved group. Moreover, we validated the findings in an independent US Latino population with distinct dietary patterns, which strengthens the generalizability of the results and reduces the likelihood of false-positive findings. Additionally, both the discovery and validation populations are well-phenotyped, allowing for adjustment of potential confounders and enhancing the robustness of the findings. Second, this study evaluated five well-recognized healthy and unhealthy dietary patterns, offering a comprehensive assessment of influences of dietary habits on the gut microbiome.

However, there are also some limitations. First, dietary data in SCCS were collected only once at baseline, and the stool samples were collected ∼13 years later. This long-time gap and possible changes in participants’ dietary habits during the period, may introduce potential misclassification and likely bias the results toward the null. Second, the use of different dietary assessment methods in SCCS (FFQ) and HCHS/SOL (24-h dietary recall) may limit the comparability and validation of findings across cohorts. Third, some microbial taxa identified in SCCS are not available for validation in HCHS/SOL, possibly due to differences in population characteristics, stool collection and processing, or microbiome sequencing platforms. Fourth, microbiome and obesity status were assessed concurrently, raising the possibility of reverse causation. Lastly, although we controlled for a wide range of covariates in the analysis, residual confounding from unmeasured variables cannot be ruled out. Future studies with large sample sizes, repeated diet and stool sample collections, and long-term follow-up are needed to prospectively investigate diet-microbiome-disease associations.

In conclusion, this two-stage study among Black/African and Hispanic/Latino Americans linked healthy dietary patterns to a lower abundance of *Collinsella* and *C. aerofaciens*, which were positively associated with obesity and mediated a significant portion of the diet-obesity association. These findings support the role of gut microbiome as a potential mediator through which diet influences metabolic health. Potential effect modifications by lifestyle factors and metabolic disease status underscore the complex nature of diet-microbiome interactions. Continued research is needed to better understand how dietary interventions can be tailored to optimize microbiome composition and reduce disease risk in diverse populations.

## Author contributions

The authors’ contributions were as follows – LW and DY: designed research; LW, YZ, and SN: analyzed data; LW: drafted the manuscript; DY: supervised study; all authors: contributed to the interpretation of results and reviewing and editing the paper and approved the final version of the manuscript. DY has full access to all the data in the study and takes responsibility for its integrity and accuracy.

## Acknowledgements

The authors thank the staff and participants of the SCCS and HCHS/SOL for their important contributions.

## Data Availability

Data described in the manuscript, code book, and analytic code will be made available upon request pending application to and approval by corresponding author and cohort committee.

## Declaration of Generative AI and AI-assisted technologies in the writing process

The author(s) declare that no generative AI or AI-assisted technologies were used in the writing of this manuscript.

## Abbreviations

BMI: body mass index;
CHC: Community Health Center;
CMD: cardiometabolic diseases;
DASH: Dietary Approaches to Stop Hypertension;
DGA: Dietary Guidelines for Americans;
EDIH: Empirical Dietary Index for Hyperinsulinemia;
EDIP: Empirical Dietary Inflammatory Potential;
FDR: false discovery rate;
FFQ: food frequency questionnaire;
HCHS/SOL: Hispanic Community Health Study / Study of Latinos;
HEI: Healthy Eating Index;
MET: metabolic equivalent of task;
SCCS: Southern Community Cohort Study;
SCFA: short-chain fatty acids;
UPF: ultra-processed foods;
USDA: U.S. Department of Agriculture.

## Reference

1. Afshin A, Sur PJ, Fay KA, Cornaby L, Ferrara G, Salama JS, Mullany EC, Abate KH, Abbafati C, Abebe Z, et al. Health effects of dietary risks in 195 countries, 1990–2017: a systematic analysis for the Global Burden of Disease Study 2017. The Lancet 2019;393:1958–72.

2. Ross FC, Patangia D, Grimaud G, Lavelle A, Dempsey EM, Ross RP, Stanton C. The interplay between diet and the gut microbiome: implications for health and disease. Nat Rev Microbiol 2024;22:671–86.

3. Singh RK, Chang H-W, Yan D, Lee KM, Ucmak D, Wong K, Abrouk M, Farahnik B, Nakamura M, Zhu TH, et al. Influence of diet on the gut microbiome and implications for human health. J Transl Med 2017;15:73.

4. David LA, Maurice CF, Carmody RN, Gootenberg DB, Button JE, Wolfe BE, Ling AV, Devlin AS, Varma Y, Fischbach MA, et al. Diet rapidly and reproducibly alters the human gut microbiome. Nature 2014;505:559–63.

5. Bolte LA, Vich Vila A, Imhann F, Collij V, Gacesa R, Peters V, Wijmenga C, Kurilshikov A, Campmans-Kuijpers MJE, Fu J, et al. Long-term dietary patterns are associated with pro-inflammatory and anti-inflammatory features of the gut microbiome. Gut 2021;70:1287–98.

6. Asnicar F, Berry SE, Valdes AM, Nguyen LH, Piccinno G, Drew DA, Leeming E, Gibson R, Le Roy C, Khatib HA, et al. Microbiome connections with host metabolism and habitual diet from 1,098 deeply phenotyped individuals. Nat Med 2021;27:321–32.

7. Maskarinec G, Hullar MAJ, Monroe KR, Shepherd JA, Hunt J, Randolph TW, Wilkens LR, Boushey CJ, Le Marchand L, Lim U, et al. Fecal Microbial Diversity and Structure Are Associated with Diet Quality in the Multiethnic Cohort Adiposity Phenotype Study. The Journal of Nutrition 2019;149:1575–84.

8. Peters BA, Xing J, Chen G-C, Usyk M, Wang Z, McClain AC, Thyagarajan B, Daviglus ML, Sotres-Alvarez D, Hu FB, et al. Healthy dietary patterns are associated with the gut microbiome in the Hispanic Community Health Study/Study of Latinos. The American Journal of Clinical Nutrition 2023;117:540–52.

9. Ma E, Maskarinec G, Lim U, Boushey CJ, Wilkens LR, Setiawan VW, Le Marchand L, Randolph TW, Jenkins IC, Curtis KR, et al. Long-term association between diet quality and characteristics of the gut microbiome in the multiethnic cohort study. Br J Nutr 2022;128:93–102.

10. Baldeon AD, McDonald D, Gonzalez A, Knight R, Holscher HD. Diet Quality and the Fecal Microbiota in Adults in the American Gut Project. The Journal of Nutrition 2023;153:2004–15.

11. Gacesa R, Kurilshikov A, Vich Vila A, Sinha T, Klaassen MAY, Bolte LA, Andreu-Sánchez S, Chen L, Collij V, Hu S, et al. The Dutch Microbiome Project defines factors that shape the healthy gut microbiome [Internet]. Microbiology; 2020 Nov. Available from: http://biorxiv.org/lookup/doi/10.1101/2020.11.27.401125

12. Breuninger TA, Wawro N, Breuninger J, Reitmeier S, Clavel T, Six-Merker J, Pestoni G, Rohrmann S, Rathmann W, Peters A, et al. Associations between habitual diet, metabolic disease, and the gut microbiota using latent Dirichlet allocation. Microbiome 2021;9:61.

13. Cotillard A, Cartier-Meheust A, Litwin NS, Chaumont S, Saccareau M, Lejzerowicz F, Tap J, Koutnikova H, Lopez DG, McDonald D, et al. A posteriori dietary patterns better explain variations of the gut microbiome than individual markers in the American Gut Project. The American Journal of Clinical Nutrition 2022;115:432–43.

14. Shi N, Nepal S, Hoobler R, Menni C, Playdon MC, Spakowicz D, Wells PM, Steves CJ, Clinton SK, Tabung FK. Pro-inflammatory and hyperinsulinaemic dietary patterns are associated with specific gut microbiome profiles: a TwinsUK cohort study. Gut Microbiome (Camb) 2024;5:e12.

15. Nepal S, Shi N, Hoyd R, Spakowicz DJ, Orwoll E, Shikany JM, Napoli N, Tabung FK. Role of insulinemic and inflammatory dietary patterns on gut microbial composition and circulating biomarkers of metabolic health among older American men. Gut Microbes 2025;17:2497400.

16. Cuevas-Sierra A, Milagro FI, Aranaz P, Martínez JA, Riezu-Boj JI. Gut Microbiota Differences According to Ultra-Processed Food Consumption in a Spanish Population. Nutrients 2021;13:2710.

17. Fernandes AE, Rosa PWL, Melo ME, Martins RCR, Santin FGO, Moura AMSH, Coelho GSMA, Sabino EC, Cercato C, Mancini MC. Differences in the gut microbiota of women according to ultra-processed food consumption. Nutrition, Metabolism and Cardiovascular Diseases 2023;33:84–9.

18. Atzeni A, Martínez MÁ, Babio N, Konstanti P, Tinahones FJ, Vioque J, Corella D, Fitó M, Vidal J, Moreno-Indias I, et al. Association between ultra-processed food consumption and gut microbiota in senior subjects with overweight/obesity and metabolic syndrome. Front Nutr 2022;9:976547.

19. Atzeni A, Hernández-Cacho A, Khoury N, Babio N, Belzer C, Vioque J, Corella D, Fitó M, Clish C, Vidal J, et al. The link between ultra-processed food consumption, fecal microbiota, and metabolomic profiles in older mediterranean adults at high cardiovascular risk. Nutr J 2025;24:62.

20. Signorello LB, Hargreaves MK, Blot WJ. The Southern Community Cohort Study: investigating health disparities. J Health Care Poor Underserved 2010;21:26–37.

21. Sorlie PD, Avilés-Santa LM, Wassertheil-Smoller S, Kaplan RC, Daviglus ML, Giachello AL, Schneiderman N, Raij L, Talavera G, Allison M, et al. Design and implementation of the Hispanic Community Health Study/Study of Latinos. Ann Epidemiol 2010;20:629–41.

22. Buchowski MS, Schlundt DG, Hargreaves MK, Hankin JH, Signorello LB, Blot WJ. Development of a culturally sensitive food frequency questionnaire for use in the Southern Community Cohort Study. Cell Mol Biol (Noisy-le-grand) 2003;49:1295–304.

23. Signorello LB, Munro HM, Buchowski MS, Schlundt DG, Cohen SS, Hargreaves MK, Blot WJ. Estimating Nutrient Intake From a Food Frequency Questionnaire: Incorporating the Elements of Race and Geographic Region. American Journal of Epidemiology 2009;170:104–11.

24. Schlundt DG, Buchowski MS, Hargreaves MK, Hankin JH, Signorello LB, Blot WJ. Separate estimates of portion size were not essential for energy and nutrient estimation: results from the Southern Community Cohort food-frequency questionnaire pilot study. Public Health Nutr 2007;10:245–51.

25. Wang L, Pan X-F, Munro HM, Shrubsole MJ, Yu D. Consumption of ultra-processed foods and all-cause and cause-specific mortality in the Southern Community Cohort Study. Clin Nutr 2023;42:1866–74.

26. Guenther PM, Casavale KO, Reedy J, Kirkpatrick SI, Hiza HAB, Kuczynski KJ, Kahle LL, Krebs-Smith SM. Update of the Healthy Eating Index: HEI-2010. J Acad Nutr Diet 2013;113:569–80.

27. Yu D, Sonderman J, Buchowski MS, McLaughlin JK, Shu X-O, Steinwandel M, Signorello LB, Zhang X, Hargreaves MK, Blot WJ, et al. Healthy Eating and Risks of Total and Cause-Specific Death among Low-Income Populations of African-Americans and Other Adults in the Southeastern United States: A Prospective Cohort Study. Stuckler D, editor. PLoS Med 2015;12:e1001830.

28. Health N, Services USDH, Institute NHLB, Lung BNHI. Your Guide to Lowering Your Blood Pressure with DASH: DASH Eating Plan [Internet]. CreateSpace Independent Publishing Platform; 2012. Available from: https://books.google.com/books?id=iQOYvwEACAAJ

29. Chang RS, Xu M, Brown SH, Cohen SS, Yu D, Akwo EA, Dixon D, Lipworth L, Gupta DK. Relation of the Dietary Approaches to Stop Hypertension Dietary Pattern to Heart Failure Risk and Socioeconomic Status (from the Southern Community Cohort Study). The American Journal of Cardiology 2022;169:71–7.

30. Tabung FK, Smith-Warner SA, Chavarro JE, Wu K, Fuchs CS, Hu FB, Chan AT, Willett WC, Giovannucci EL. Development and Validation of an Empirical Dietary Inflammatory Index. J Nutr 2016;146:1560–70.

31. Tabung FK, Wang W, Fung TT, Hu FB, Smith-Warner SA, Chavarro JE, Fuchs CS, Willett WC, Giovannucci EL. Development and validation of empirical indices to assess the insulinaemic potential of diet and lifestyle. Br J Nutr 2016;116:1787–98.

32. Monteiro CA, Cannon G, Levy R, Moubarac J-C, Jaime P, Martins AP, Canella D, Louzada M, Parra D. NOVA. The star shines bright. World Nutrition 2016;7:28–38.

33. Liu L, Nguyen SM, Wang L, Shi J, Long J, Cai Q, Shrubsole MJ, Shu X-O, Zheng W, Yu D. Associations of alcohol intake with gut microbiome: a prospective study in a predominantly low-income Black/African American population. The American Journal of Clinical Nutrition 2025;121:134–40.

34. Sinha R, Chen J, Amir A, Vogtmann E, Shi J, Inman KS, Flores R, Sampson J, Knight R, Chia N. Collecting Fecal Samples for Microbiome Analyses in Epidemiology Studies. Cancer Epidemiol Biomarkers Prev 2016;25:407–16.

35. Blanco-Míguez A, Beghini F, Cumbo F, McIver LJ, Thompson KN, Zolfo M, Manghi P, Dubois L, Huang KD, Thomas AM, et al. Extending and improving metagenomic taxonomic profiling with uncharacterized species using MetaPhlAn 4. Nat Biotechnol 2023;41:1633–44.

36. Abubucker S, Segata N, Goll J, Schubert AM, Izard J, Cantarel BL, Rodriguez-Mueller B, Zucker J, Thiagarajan M, Henrissat B, et al. Metabolic Reconstruction for Metagenomic Data and Its Application to the Human Microbiome. Eisen JA, editor. PLoS Comput Biol 2012;8:e1002358.

37. Suzek BE, Wang Y, Huang H, McGarvey PB, Wu CH, UniProt Consortium. UniRef clusters: a comprehensive and scalable alternative for improving sequence similarity searches. Bioinformatics 2015;31:926–32.

38. Caspi R, Billington R, Ferrer L, Foerster H, Fulcher CA, Keseler IM, Kothari A, Krummenacker M, Latendresse M, Mueller LA, et al. The MetaCyc database of metabolic pathways and enzymes and the BioCyc collection of pathway/genome databases. Nucleic Acids Res 2016;44:D471–480.

39. Bowyer RCE, Jackson MA, Pallister T, Skinner J, Spector TD, Welch AA, Steves CJ. Use of dietary indices to control for diet in human gut microbiota studies. Microbiome 2018;6:77.

40. Ericson U, Brunkwall L, Hellstrand S, Nilsson PM, Orho-Melander M. A Health-Conscious Food Pattern Is Associated with Prediabetes and Gut Microbiota in the Malmö Offspring Study. J Nutr 2020;150:861–72.

41. Iqbal M, Yu Q, Tang J, Xiang J. Unraveling the gut microbiota’s role in obesity: key metabolites, microbial species, and therapeutic insights. Kendall MM, editor. J Bacteriol 2025;e00479–24.

42. Frost F, Storck LJ, Kacprowski T, Gärtner S, Rühlemann M, Bang C, Franke A, Völker U, Aghdassi AA, Steveling A, et al. A structured weight loss program increases gut microbiota phylogenetic diversity and reduces levels of Collinsella in obese type 2 diabetics: A pilot study. Taheri S, editor. PLoS ONE 2019;14:e0219489.

43. Companys J, Gosalbes MJ, Pla-Pagà L, Calderón-Pérez L, Llauradó E, Pedret A, Valls RM, Jiménez-Hernández N, Sandoval-Ramirez BA, Del Bas JM, et al. Gut Microbiota Profile and Its Association with Clinical Variables and Dietary Intake in Overweight/Obese and Lean Subjects: A Cross-Sectional Study. Nutrients 2021;13:2032.

44. van Soest APM, Hermes GDA, Berendsen AAM, van de Rest O, Zoetendal EG, Fuentes S, Santoro A, Franceschi C, de Groot LCPGM, de Vos WM. Associations between Pro-and Anti-Inflammatory Gastro-Intestinal Microbiota, Diet, and Cognitive Functioning in Dutch Healthy Older Adults: The NU-AGE Study. Nutrients 2020;12:3471.

45. Astbury S, Atallah E, Vijay A, Aithal GP, Grove JI, Valdes AM. Lower gut microbiome diversity and higher abundance of proinflammatory genus Collinsella are associated with biopsy-proven nonalcoholic steatohepatitis. Gut Microbes 2020;11:569–80.

46. Gomez-Arango LF, Barrett HL, Wilkinson SA, Callaway LK, McIntyre HD, Morrison M, Dekker Nitert M. Low dietary fiber intake increases Collinsella abundance in the gut microbiota of overweight and obese pregnant women. Gut Microbes 2018;9:189–201.

47. Candela M, Biagi E, Soverini M, Consolandi C, Quercia S, Severgnini M, Peano C, Turroni S, Rampelli S, Pozzilli P, et al. Modulation of gut microbiota dysbioses in type 2 diabetic patients by macrobiotic Ma-Pi 2 diet. Br J Nutr 2016;116:80–93.

48. Song AA-L, In LLA, Lim SHE, Rahim RA. A review on Lactococcus lactis: from food to factory. Microb Cell Fact 2017;16:55.

49. Song YL, Liu CX, McTeague M, Summanen P, Finegold SM. Clostridium bartlettii sp. nov., isolated from human faeces. Anaerobe 2004;10:179–84.

50. Cunningham AL, Stephens JW, Harris DA. Gut microbiota influence in type 2 diabetes mellitus (T2DM). Gut Pathog 2021;13:50.

