## Supplementary file for "Discovery and Validation of Gut Microbiome Features Associated with Dietary Patterns in U.S. Black/African and Hispanic/Latino Populations"

**Supplementary Table 1. Components of EDIP and EDIH and included food items in the SCCS and HCHS/SOL**

| Components | SCCS<br>Included FFQ Variables | HCHS/SOL<br>Included Food names | Weights |
|---|---|---|---|
| **EDIP** | | | |
| Processed meat | LunchMeats, BaconSausage, HotDogSausage | Cured (lean) pork, (lean) cold cuts and sausage | 165.03 |
| Red meat | FriedBeef, OtherGroundBeef, BeefMixedDishes, Beef, PorkHam, Hamburger | (Lean) beef, (lean) veal, (lean) lamb, (lean) fresh pork, game | 140.19 |
| Organ meat | OrganMeats | Organ meats | 144.61 |
| Other fish | Tuna, FriedSeafood, Seafood | Lean fish, fried fish, (fried) shellfish | 252.45 |
| Other vegetables | OtherVeg, Corn | Other vegetables, fried vegetables, vegetable juice, and pickled foods | 136.14 |
| Refined grains | WhiteBreads, Biscuits, DoughnutPastry, Rice, Pasta | Refined grain | 81.21 |
| High-energy beverages | RegSoda, SweetenedFruitDrinks | Sweetened fruit drinks, soft drinks | 156.85 |
| Low-energy beverages | DietSoda | Artificially sweetened/ unsweetened soft drinks, artificially sweetened fruit drinks | 94.77 |
| Tomatoes | Tomatoes | Tomato | 167.92 |
| Beer | LightBeer, RegBeer | Beer and ale | -136.99 |
| Wine | WhiteWine, RedWine | Wine | -249.70 |
| Tea | Tea | Sweetened, artificially sweetened, and unsweetened tea | -42.25 |
| Coffee | RegCoffee, DecafCoffee | Sweetened, artificially sweetened, and unsweetened coffee | -83.18 |
| Dark yellow vegetables | Carrots, SweetPotatoes | Deep-yellow vegetables | -165.37 |
| Leafy green vegetables | Greens, Lettuce | Dark-green Vegetables | -190.29 |
| Snacks | SaltySnacks, Popcorn, CrackersPretzels | Snack chips, popcorn, crackers, fruit- and vegetable-based savory snack | -45.08 |
| Fruit juice | FruitJuice | Citrus and non-citrus juice | -58.95 |
| Pizza | Pizza | Pizza | -1175.21 |

| EDIH | | | |
|---|---|---|---|
| Red meat | FriedBeef, OtherGroundBeef, BeefMixedDishes, Beef, PorkHam, Hamburger | (Lean) beef, (lean) veal, (lean) lamb, (lean) fresh pork, game | 0.250 |
| Low-energy beverage | DietSoda | Artificially sweetened/ unsweetened soft drinks, artificially sweetened fruit drinks | 0.053 |
| Cream soups | No such question in the FFQ for cream soups. This component is set to missing for all participants. | Same as SCCS. | 0.787 |
| Processed meat | LunchMeats, BaconSausage, HotDogSausage | Cured (lean) pork, (lean) cold cuts and sausage | 0.199 |
| Margarine | Margarine | Regular/reduced fat margarine | 0.054 |
| Poultry | FriedChicken, ChickenTurkey, ChickenMixedDishes | (Lean) poultry and fried chicken | 0.183 |
| Butter | Butter | Regular/reduced fat butter and other animal fats | 0.094 |
| French fries | FriedPotatoes | Fried potato | 0.581 |
| Other fish | Tuna, FriedSeafood, Seafood | Lean fish, fried fish, (fried) shellfish | 0.172 |
| High-energy beverages | RegSoda, SweetenedFruitDrinks | Sweetened fruit drinks, soft drinks | 0.104 |
| Tomatoes | Tomatoes | Tomato | 0.095 |
| Low-fat dairy | SkimMilk, LowFatMilk, FroYoSherbet, CottageCheeseYogurt | Reduced/Low fat/fat free milk, ready-to-drink flavored milk; Low fat/fat free cheese, (artificially) un/sweetened yogurt, cream; (artificially) un/sweetened flavored milk beverage powder with non-fat dry milk | 0.025 |
| Eggs | Eggs | Egg (substitute) | 0.124 |
| Wine | WhiteWine, RedWine | Wine | -0.165 |
| Coffee | RegCoffee, DecafCoffee | Sweetened, artificially sweetened, and unsweetened coffee | -0.035 |
| Whole fruits | Bananas, ApplesPears, Melon, GrapesBerries, Citrus, Peaches, OtherFruit | Citrus and non-citrus fruit, avocado and similar | -0.029 |

| | | | |
|---|---|---|---|
| High-fat dairy | WholeMilk, Cream, Cheese, IceCream | Full fat milk, ready-to-drink flavored milk; full/reduced fat cheese, and cream; (artificially) un/sweetened whole milk yogurt | -0.046 |
| Leafy green vegetables | Greens, Lettuce | Dark-green Vegetables | -0.055 |

Abbreviations: EDIH, empirical dietary index for hyperinsulinemia; EDIP, empirical dietary inflammatory pattern; FFQ, food frequency questionnaire.

**Supplementary Table 2. Baseline characteristics of included participants in the SCCS and HCHS/SOL**

| Baseline characteristics | SCCS (N=514) | HCHS/SOL (N=2,133) |
|---|---|---|
| **Enrollment age, years** | 53.2 ± 7.7 | 50.8 ± 11.1 |
| **Age at stool collection, years** | 66.5 (7.8) | 58.1 (11.2) |
| **Time interval from enrollment to stool collection, years** | 13.3 (2.0) | 7.3 (1.0) |
| **Sex, n (%)** | | |
| Male | 144 (28.0) | 797 (37.4) |
| Female | 370 (72.0) | 1336 (62.6) |
| **Educational attainment, n (%)** | | |
| Less than high school | 83 (16.2) | 805 (37.7) |
| High school or equivalent | 178 (34.6) | 496 (23.3) |
| More than high school | 253 (49.2) | 832 (39.0) |
| **Smoking status, n (%)** | | |
| Never | 243 (47.3) | 418 (19.6) |
| Former | 133 (25.8) | 763 (35.8) |
| Current | 138 (26.9) | 952 (44.6) |
| **Alcohol drinking, n (%)** | | |
| Never | 259 (50.4) | 1340 (62.8) |
| Former/ Current | 255 (49.6) | 793 (37.2) |
| **Total physical activity, MET-hours/day** | 3.1 ± 2.4 | 9.5 ± 15.0 |
| **Total sitting time, hours/day** | 9.6 ± 4.4 | 3.8 ± 3.0 |
| **Total energy intake, kcal/day** | 2437 ± 1358 | 1787 ± 675 |
| **BMI, kg/m$^2$** | 30.9 ± 6.8 | 29.9 ± 5.8 |
| **Prevalent medical conditions, n (%)** | | |
| Cancer | 34 (6.6) | 77 (3.6) |
| CVD | 36 (7.0) | 204 (9.6) |
| Diabetes | 80 (15.6) | 543 (25.5) |
| Hypertension | 286 (55.6) | 816 (38.3) |
| Hypercholesterolemia | 200 (38.9) | 1073 (50.3) |
| **Dietary scores** | | |
| HEI | 60.7 ± 12.9 | 61.3 ± 14.0 |
| DASH | 48.9 ± 10.0 | 44.8 ± 10.6 |
| EDIP | -0.16 ± 0.88 | 1.04 ± 0.77 |
| EDIH | 0.2 ± 0.2 | 1.24 ± 0.83 |

*Data was presented as mean ± standard deviation for continuous variables and number (percentage) for categorical variables

**Supplementary Figure 1. Correlations between HEI, DASH, EDIP, EDIH, and UPF in the SCCS**

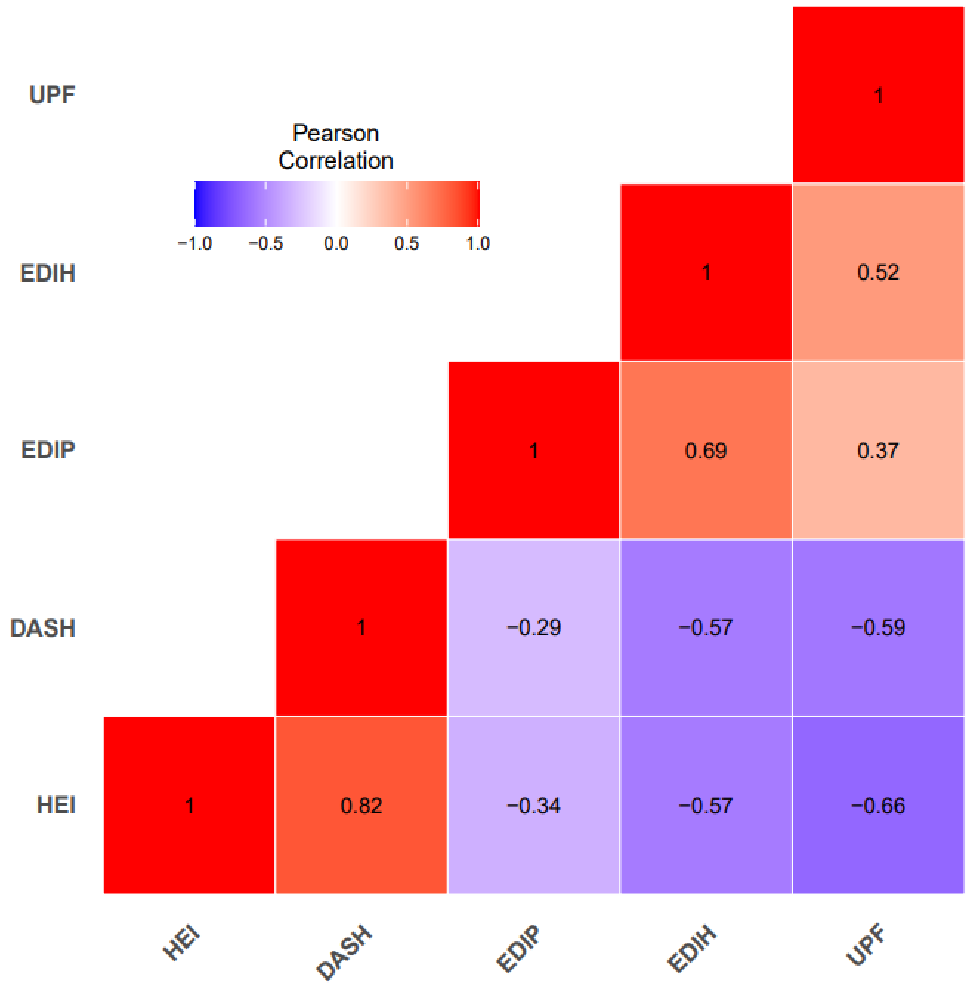

**Supplementary Figure 2. Differences in Shannon index across tertiles of dietary patterns in the SCCS**

Adjusted for enrollment age, time interval between enrollment and stool collection, sex, enrollment source, education, income, smoking status, pack years, number of alcohol drinks per day, physical activity, sitting hours, total daily calories, BMI, and disease history at baseline including cancer, CVD (CHD and stroke), diabetes, hypertension, and dyslipidemia.

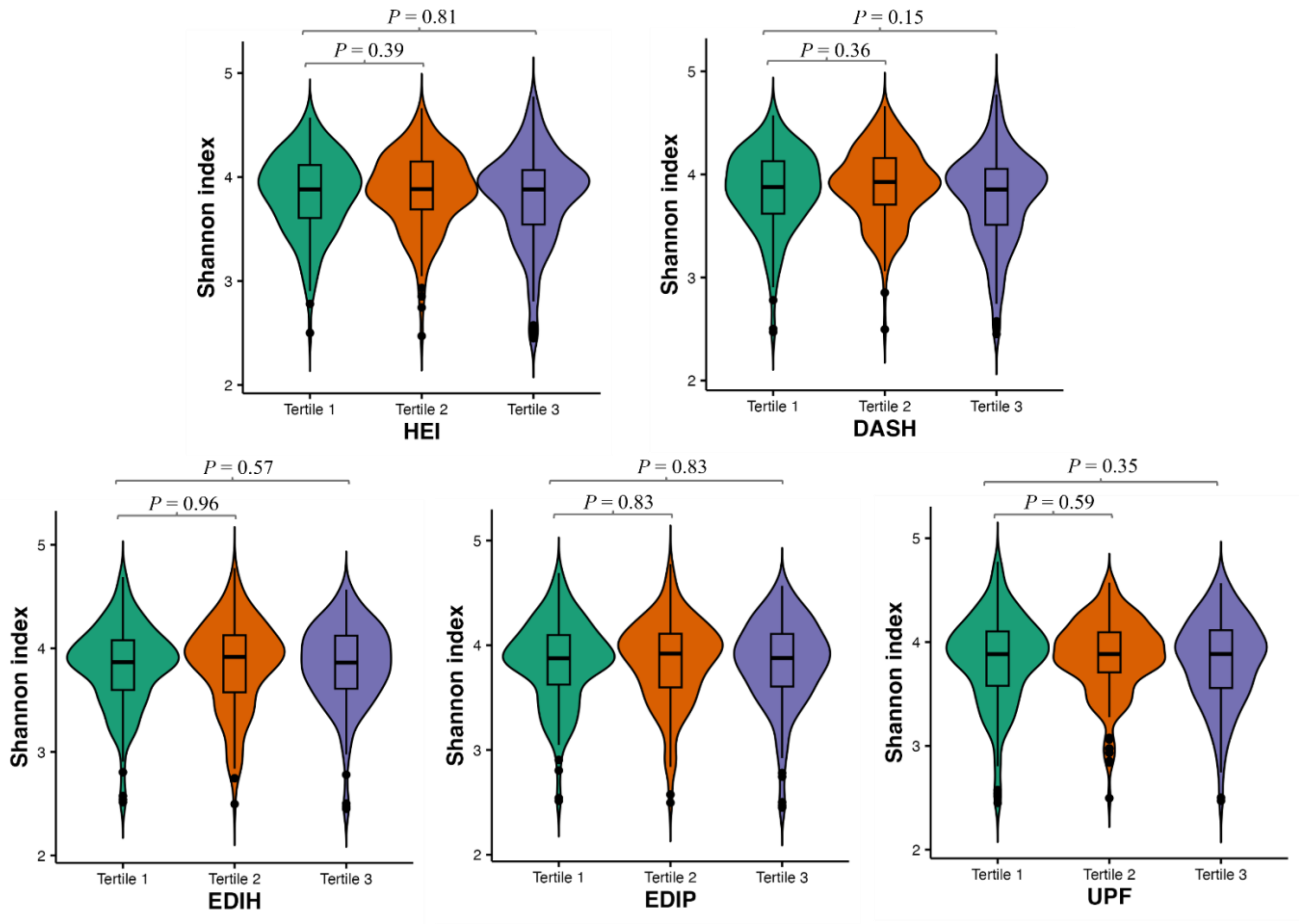

**Supplementary Figure 3. Proportion of variation in Bray–Curtis dissimilarity explained by dietary patterns in the SCCS**

Adjusted for enrollment age, time interval between enrollment and stool collection, sex, enrollment source, education, income, smoking status, pack years, number of alcohol drinks per day, physical activity, sitting hours, total daily calories, BMI, and disease history at baseline including cancer, CVD (CHD and stroke), diabetes, hypertension, and dyslipidemia.

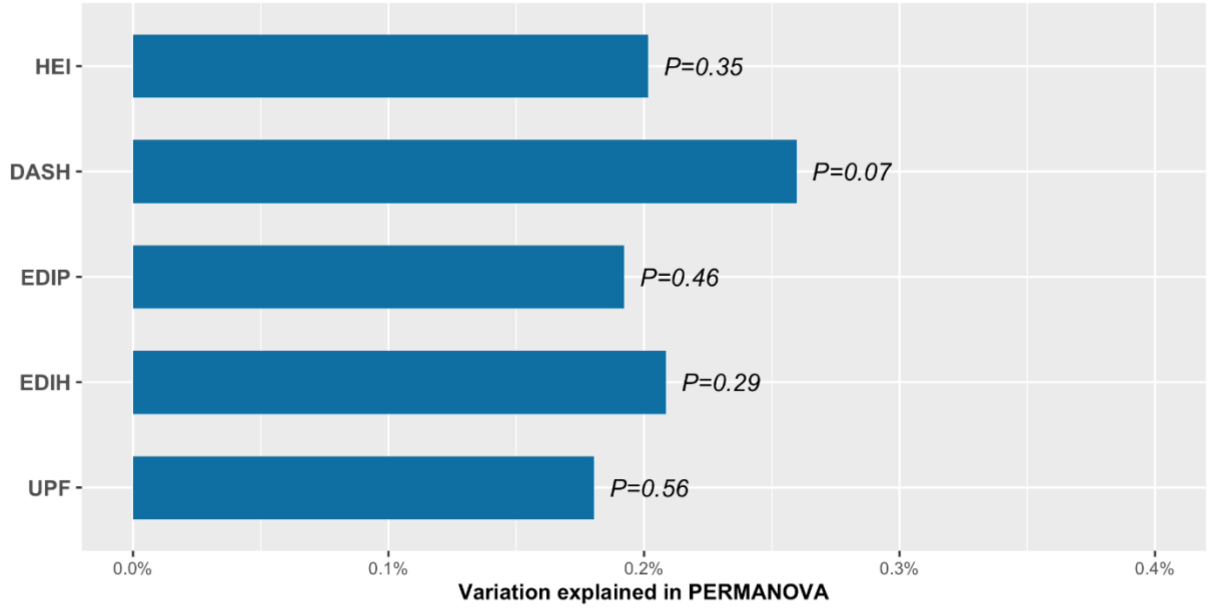

**Supplementary Figure 4. Mediation effects of gut microbial taxa on diet and obesity in the SCCS**

In the mediation analysis, covariates adjusted in the model included enrollment age, time interval between enrollment and stool collection, sex, enrollment source, education, income, smoking status, pack years, number of alcohol drinks per day, physical activity, sitting hours, total daily calories, baseline BMI, and disease history at baseline including cancer, CVD, diabetes, hypertension, and hypercholesterolemia.

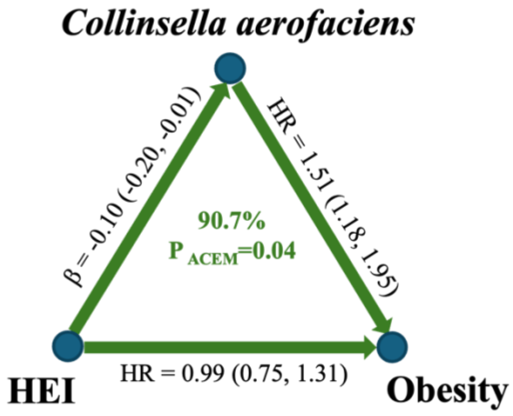


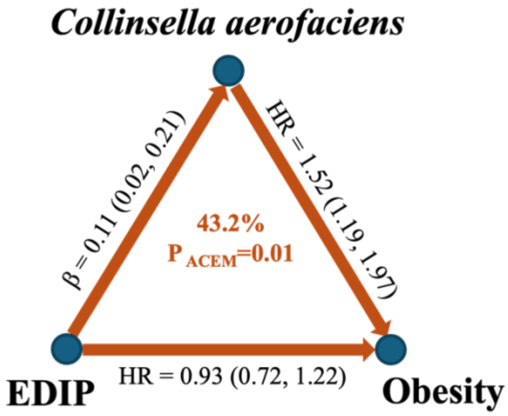
